# MFDNN: Multi-channel feature deep neural network algorithm to identify Covid19 chest X-ray images

**DOI:** 10.1101/2021.08.04.21261235

**Authors:** Liangrui Pan, Boya Ji, Xiaoqi Wang, Shaolaing Peng

## Abstract

The use of chest X-ray images (CXI) to detect Severe Acute Respiratory Syndrome Coronavirus 2 (SARS CoV-2) caused by Coronavirus Disease 2019 (COVID-19) is life-saving important for both patients and doctors. This research proposed a multi-channel feature deep neural network algorithm to screen people infected with COVID-19. The algorithm integrates data oversampling technology and a multi-channel feature deep neural network model to carry out the training process in an end-to-end manner. In the experiment, we used a publicly available CXI database with 10,192 Normal, 6012 Lung Opacity (Non-COVID lung infection), and 1345 Viral Pneumonia images. Compared with traditional deep learning models (Densenet201, ResNet50, VGG19, GoogLeNet), the MFDNN model obtains an average test accuracy of 93.19% in all data. Furthermore, in each type of screening, the precision, recall, and F1 Score of the MFDNN model are also better than traditional deep learning networks. Secondly, compared with the latest CoroDet model, the MFDNN algorithm is 1.91% higher than the CoroDet model in the experiment of detecting the four categories of COVID19 infected persons. Finally, our experimental code will be placed at https://github.com/panliangrui/covid19.

## 1. Introduction

SARS-CoV-2 causes COVID-19. Since the first report, it has become a global pandemic, with 180 million confirmed cases and 3.91 million deaths. It is extremely contagious characteristics and delayed vaccination has made developing countries vulnerable to virus attacks. Now the nucleic acid detection mechanism has played an essential role in screening the flow of people. Reverse Transcription Polymerase Chain Reaction (RT-PCR) is considered to be the most standard diagnostic technology available[1]. However, its sensitivity is relatively low, and the result is highly dependent on the sample area obtained and heavily reliant on the operator’s technique [2]. The important thing is that this method takes time. However, time is a key factor in isolating, preventing and treating people infected with COVID-19, which will limit the efficiency of COVID-19 screening. With the global spread of COVID-19, medical research has found that CXI can identify people infected with COVID-19. Therefore, as a supplement to RT-PCR technology, it plays an essential role in detecting and evaluating people infected with COVID-19.

Computed tomography (CT), lung ultrasound (LUS), and Chest-X ray radiography are among the most commonly used imaging modalities to identify COVID-19 infections [3–5]. Because of the safety, painlessness, non-invasiveness, clear image, high-density resolution, and apparent morbidity of CXI, it is widely used in large hospitals. In addition, experienced doctors can make real-time diagnoses through CXI. Therefore, CXI is one of the most commonly used and readily available methods to detect COVID-19 infections [6]. However, there are many similarities in the CXI characteristics of patients with COVID19 and common pneumonia, which poses a huge challenge to radiologists in diagnosing patients with COVID19.

In recent years, artificial intelligence has prompted tremendous progress in the field of biomedicine, such as intelligent medical diagnosis, intelligent image recognition, intelligent health management, intelligent drug development, and medical robots [7–9]. Machine learning-based methods have developed many applications in the accurate analysis of CXI, such as diagnosing and evaluating people infected with COVID-19 [10][11]. Common machine learning algorithms include linear regression, random forest (RF), K-nearest neighbour (KNN), decision tree (DT), etc[12,13]. Abolfazl et al. used dimensionality reduction methods to extract the best features of CRX images to build an efficient machine learning classifier, and the classifier distinguishes covid-19 and non-covid-19 cases with high accuracy and sensitivity [6]. Dan et al. used three different machine learning models to predict the deterioration of the patient’s condition, compare them with the currently recommended predictors and APACHEII risk prediction scores, and obtain high sensitivity, specificity, and accuracy [14]. Mohamed et al. used the new fractional multi-channel exponential moments (FrMEMs) to extract features from CXI[15].

Then, the improved Manta-Ray Foraging Optimization (MRFO) method was used for feature selection, and the KNN method was used to classify the two types of CRX Image [15].However, deep learning is the hottest research direction in the field of machine learning. The CXR deep learning method for COVID-19 classification has been actively explored. Linda et al. proposed a deep convolutional neural network called COVID-Net to help clinicians improve screening [16]. Ali et al. proposed five models based on pre-trained convolutional neural networks (ResNet50, ResNet101, ResNet152, InceptionV3, and Inception-ResNetV2) to implement four different binary classifications: COVID-19, normal (healthy), and viral pneumonia and bacterial pneumonia) CXI has achieved a high accuracy rate [17]. Loannis et al. automatically detected CXI based on the transfer learning method of convolutional neural network and achieve 96.78%, 98.66%, and 96.46% accuracy, sensitivity, and specificity[18]. Ezz et al. proposed the COVIDX-Net network based on seven deep convolutional network models with different architectures and obtained F1 scores of 0.89 and 0.91, respectively [19].

Inspired by machine learning and deep learning and the accumulation of previous work experience, in this article, we will further explore the impact of experimental data and deep convolutional neural networks on the detection algorithm or detection system. However, in most databases, unbalanced label classes often occur, which will cause the convolutional neural network to be biased to identify image data with many class labels correctly. Therefore, the main focus of this article is to solve the accuracy of the COVID-19 detection algorithm. Around this problem, we will solve the following problems separately: (1) Deal with the imbalance of sample labels. (2) Optimize the feature extraction of the deep neural network algorithm. (3) Evaluate the classification effect of the network algorithm.

To achieve this goal, we first analyze the degree of imbalance in the sample data. We found that the amount of chest X-ray data of people who were not infected with COVID-19 was significantly more than other categories through the data set analysis. To be able to classify the chest radiograph data set more accurately, our main contributions are as follows:

1. To balance the impact of the unbalanced label data set on model training, when processing CXI, we embed the oversampling method into the end-to-end model to balance all categories of data.
2. We propose a multi-channel feature deep neural network (MFDNN) algorithm based on multi-channel input, single-channel output, and weight centralized sharing. The algorithm model concentrates the feature maps of chest radiographs from multiple channels and optimizes the feature extraction process, which simplifies classification.
3. Finally, the proposed method is compared with the classic deep neural networks VGG19, Resnet50, Desnet201. Secondly, we also compare with the latest CoroDet model. The detection accuracy of our proposed MFDNN algorithm is 1.91% higher than that of the CoroDet model.

## 2. Methods

In this section, we first introduce the flow chart of the MFDNN algorithm. It mainly includes two parts: oversampling and the MFDNN model. The first part is primarily data preprocessing, and the second part primarily uses the MFDNN model for feature extraction and patient diagnosis. Figure 1. shows our proposed algorithm classification process.

**Figure 1.**
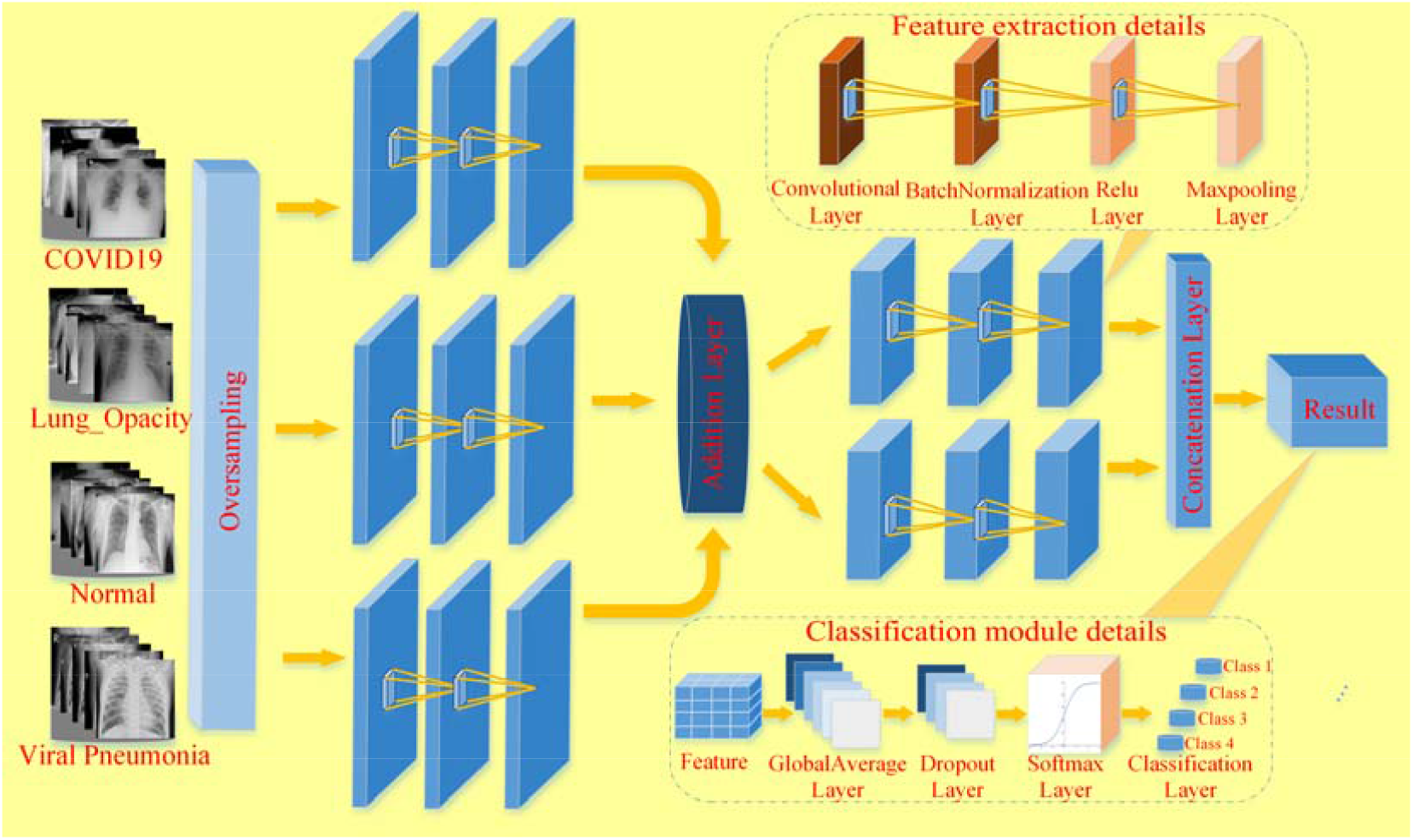
End-to-end MFDNN algorithm flow chart. It includes two parts, namely data oversampling, feature extraction and classification.

### 2.1. Oversampling

Before the experiment, we need to have a preliminary understanding of the COVID-19 database. As shown in Figure 2. (a), by comparing the characteristics of the database, the chest radiographs of people infected with COVID19 are significantly less than those of normal people. However, the steps of neural networks and human brain extraction are similar. When the probability of memorizing normal chest radiographs of the MFDNN model is greater than remembering the chest radiographs of COVID19 infected persons, the model is more likely to recognize the chest radiographs of normal people, which may lead to COVID19 infection. Therefore, the probability of screening by the user is reduced, and the model cannot be applied to the actual detection process. The experiment chooses to oversample to generate new samples for a few categories to ensure that the model has the same probability of remembering different chest radiographs. Figure 2. (b) shows the result of oversampling. The sample data size of the minority class is the same as the sample data size of the majority class.

**Figure 2.**
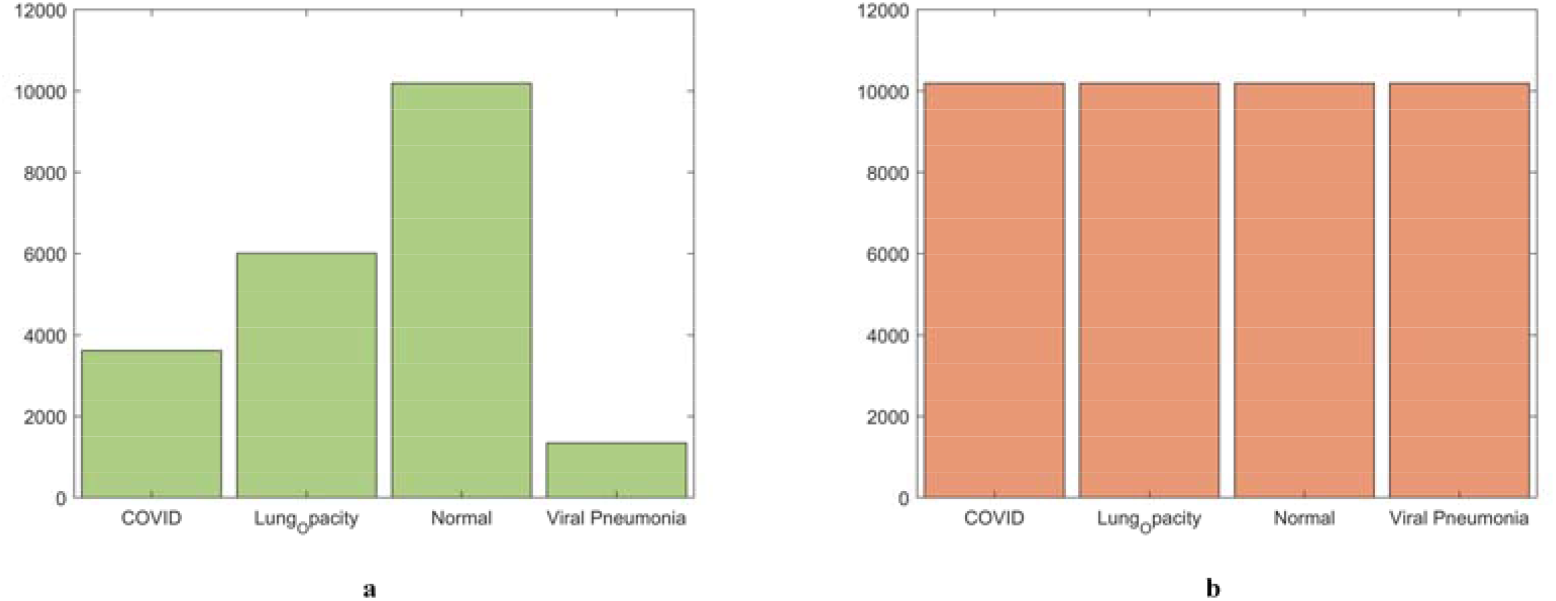
(a). The sample size of the original dataset. (b). The number of samples after oversampling.

### 2.2. Multi-channel feature deep neural network(MFDNN)

The MFDNN algorithm is experimentally designed. The oversampling dataset passes through three identical feature extraction modules. The features will be merged in the middle of the frame. Then further feature extraction is performed on the collected image features through the Siamese network, which improves image feature extraction work efficiency. We will introduce the function of each layer in detail from the feature extraction module.

The image can be seen as a high-order matrix composed of feature vectors. In feature extraction, the use of a small convolution kernel can reduce the convolution operation’s error, so the 3*3 convolution kernel is selected in the convolution layer, and the step size is 1 for matrix operation. The convolutional layer is defined as:

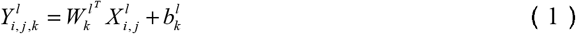

The feature value at the position (*i, j*) of the *k*-th feature map of the *l*-th layer is 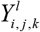 Where 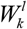 is the weight of the *l*-th layer, 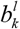 is the bias of the *l*-th layer, and 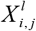 is the (*i, j*) unknown input block of the *l*-th layer. 64 convolution kernels simultaneously perform local perception and share parameters on the input image.

Batch Normalization(BN)is widely used training technique in deep networks. A BN layer whitens activations within a mini-batch of N examples for each channel dimension and transforms the whitened activations using affine parameters *γ* and *β*, Denoting by *χ* ∈ *R*^*H*×*W*×*N*^ activations each channel, BN is expressed as[20]:

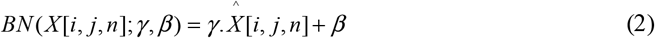

Where

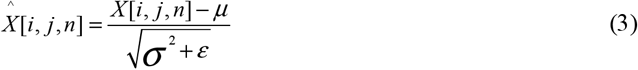

The mean and variance of activations within a mini-batch,

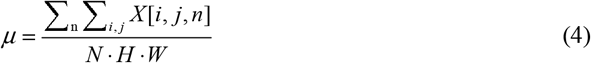

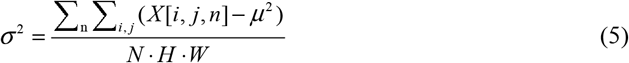

Select the Rectified Linear Unit (ReLU) function as the convolutional layer’s activation function to reduce the probability of model overfitting[21]. The ReLU function will make part of the neuron output 0 to enhance the sparsity of the network. Besides, it reduces the interdependence between parameters and alleviates the problem of overfitting. For the MFDNN model, the ReLU function enables each neuron to exert the greatest screening effect, saving a lot of calculations in the whole process, which is defined as:

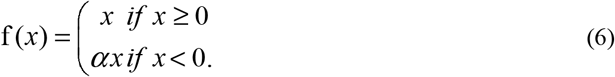

In the pooling layer, the maximum pooling method is selected to obtain the maximum value of the feature tiles from the convolutional layer as the output. The down-sampling convolution kernel size is set to 2*2, the stride size is set to 2, and the feature matrix after the pooling operation is filled in a “same” manner to alleviate the excessive sensitivity of the convolution layer to position. The maximum pooling layer reduces the parameters by reducing the dimension, removing redundant features, simplifying the network’s complexity, and other methods to achieve nonlinear feature extraction. The input 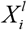 of the *l* th layer is mapped to the output through 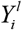 the neuron, which is defined as:

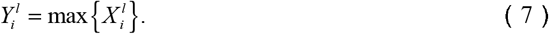

The additional layer serves as an intermediate hub for merging the output from the pooling layer, combining feature weights. Assuming the output is, its effect can be expressed as:

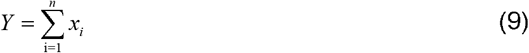

As the input *Y* of the global average pooling (GAP) layer, the learned “distribution feature representation” is selectively mapped to the labeled sample space. The activation function of each neuron in the GAP layer generally uses the ReLU function[22]. It can replace the fully connected layer in the traditional structure, thereby reducing the amount of storage required for the large weight matrix of the fully connected layer. Secondly, it also has the features and capabilities of easy fine-tuning of a pre-trained model with a conventional structure. Since the working principle of the fully connected layer involves calculating the inner product of the input vector and the weight of each row, the row size of the weight matrix needs to be the same as the number of input elements[23]. Therefore, as the input changes, we also need to adjust the weight matrix f, *W*, to a corresponding size by

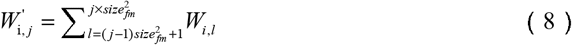

Where size _*fm*_ is the size of the input feature map, i, j is the index of the output neurons and input feature maps, and *W* ^‘^ is the modified weight matrix[23]. In view of the computational complexity in the GAP layer, the dropout layer chooses a 40% random probability to discard some feature weights to reduce the model complexity and prevent overfitting. Finally, it is classified by Softmax, and the output of multiple neurons is mapped to the interval of (0,1), which is defined as:

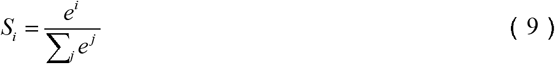

We use {(*x*^(1)^,*y*^(1)^),(*x*^(2)^,*y*^(2)^),…(*x*^(*m*)^,*y*^(*m*)^)} to represent *m* training samples and *y*^(*i*)^ to represent the label of *i* samples. The neural network is trained using gradient descent. In this article, the cross-entropy function is used to calculate the loss of the MFDNN model. For a single example, the cross-entropy loss function can be expressed as:

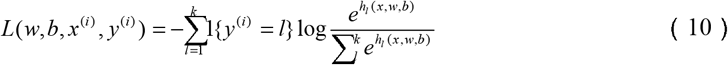

*h*_*l*_ (*x, w, b*) represents the *s* th neuron in the output layer corresponding to the *s* th type, 1{.} is the indicator function. The weight parameter is continuously updated through the backpropagation loss function. We propose an MFDNN classification algorithm for detecting COVID19 patients.

#### Algorithm 1: MFDNN Classification Model

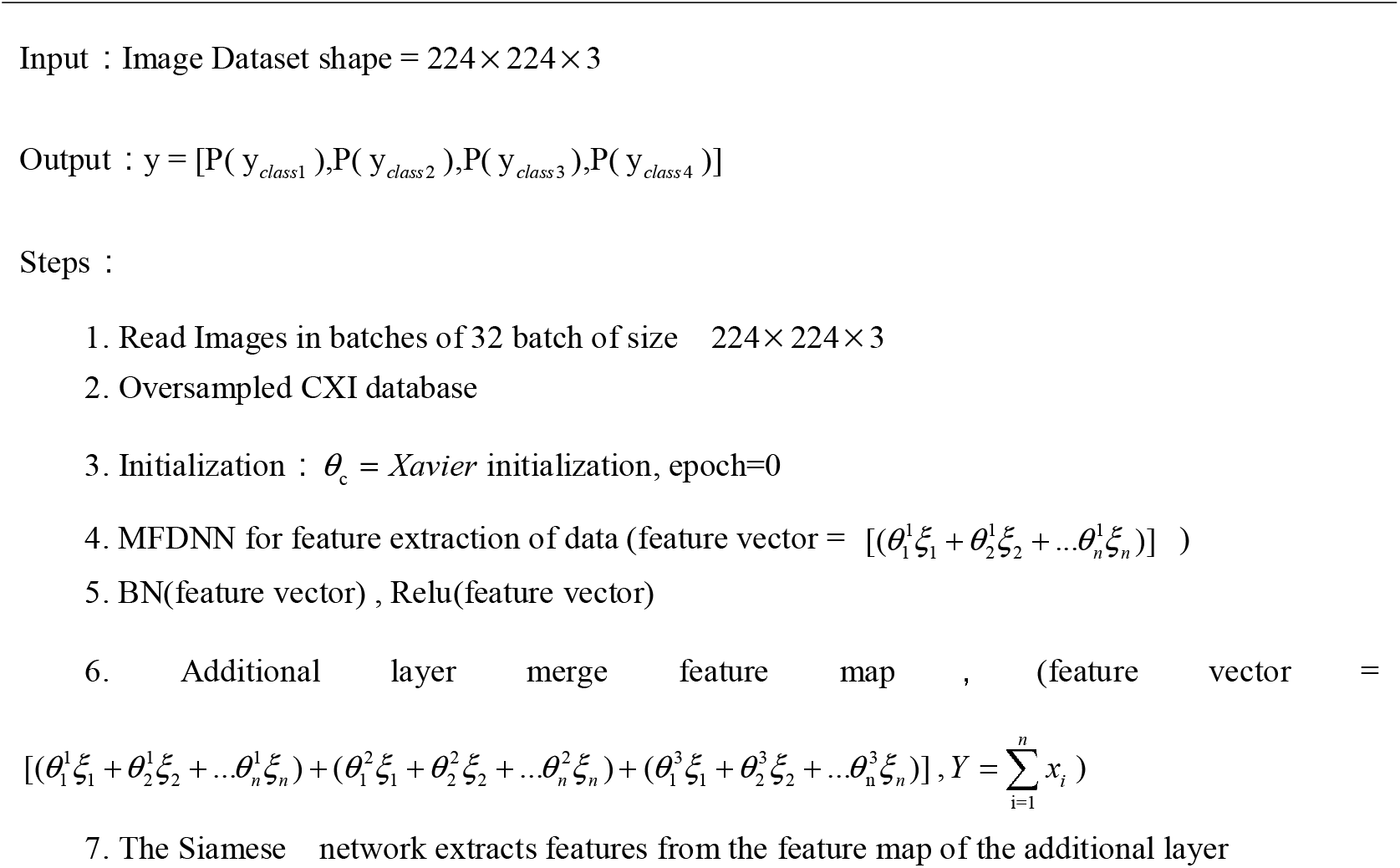
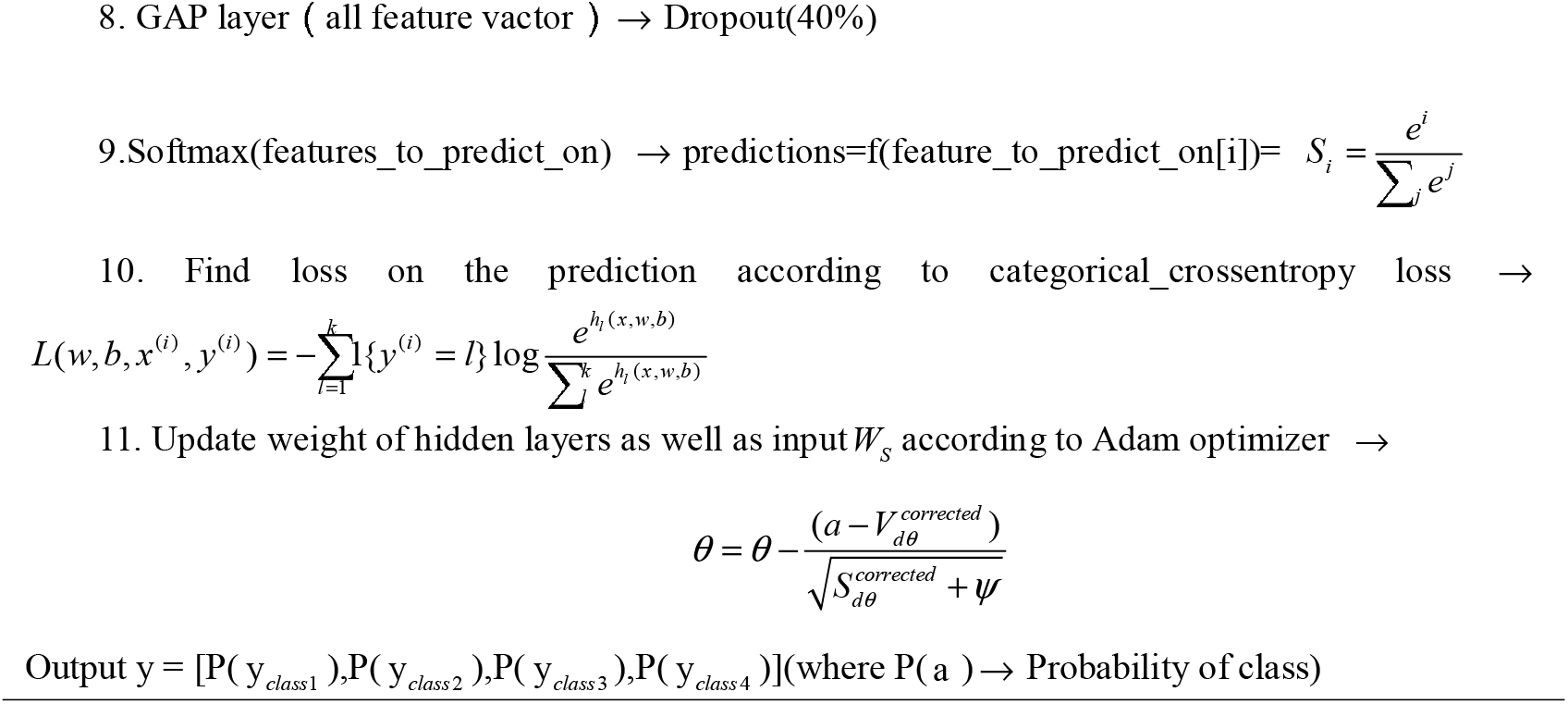

## 3. Experiments

### 3.1. Datasets

The proposed MFDNN model is trained and tested on a public dataset (Covid19 chest X-ray dataset). The data set consists of 3616 COVID-19 positive cases, 10,192 normal, 6012 lung opaque (non-COVID lung infection) and 1345 viral pneumonia images [24,25]. The dataset can be downloaded from the website (\https://www.kaggle.com/tawsifurrahman/covid19-radiography-database).

### 3.2. Experimental settings

Divide the oversampled data into the data set according to the ratio of training data and test data of 0.8:0.2, where the training data is divided into training set and validation set according to the ratio of 0.8:0.2. Before training the MFDNN model, we choose to flip the data and augment the data with translation to expand the training data to avoid over-fitting the model. The experiment is set to 30 epochs, the batch size is set to 32, and the Adam algorithm is used as the optimizer of the model. The initial learning rate is 0.003, and after each epoch, the learning rate will drop by half. Before each epoch training, the training data and verification data will be randomly shuffled. Each model is trained on a single RTX3060.

## 4. Results and discussion

### 4.1. Results

In this section, we will explain the evaluation indicators used to quantify model classification. To this end, we use an indicator based on a confusion matrix. These indicators include test accuracy, precision, recall, and F1 Score. To evaluate the model, we need to perform a detailed analysis of each category. Therefore, we need to count true positives, false positives, true negatives, and false positives[26].

1. Test accuracy: the proportion of samples correctly predicted to the total samples

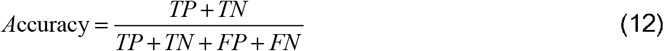
2. Precision: the ratio of true positive predictions to total positive predictions

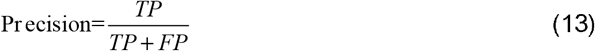
3. Recall: Ratio of true positive to the total observation made by the proposed model

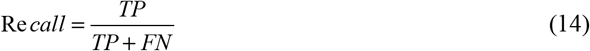
4. F1 Score: It is the harmonic mean of precision and recall

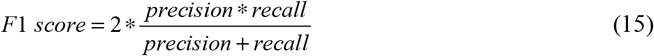
5. Confusion matrix: It is the measurement of the performance of the model. It compares the actual and predicted values in form of True Positive, False Negative, True Negative and False Positive

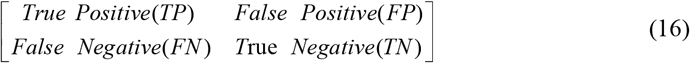

. True Positive (TP): True positive are the forecasts which were at first positive and, additionally, anticipated by the AI model as positive.
. False Positive (FP): False positives are the forecasts which were initially negative and anticipated by the AI model as positive.
. True Negative (TN): True negatives are the forecasts which were initially negative and anticipated by the AI model as unfavourable.
. False Negative (FN): False-negative are the forecasts which were initially positives and anticipated by the model as negative

The experiment first trained five models under the algorithm of MFDNN, namely Densenet201, ResNet50, VGG19, GoogLeNet, and MFDNN. Among them, the accuracy of the MFDNN model is 93.19%. Among them, the COVID category received a Recall of 0.9447 and an F1 score of 0.9358; the Lung_Opacity category received a precision of 0.9144 and an F1 score of 0.9106; the Normal class received a recall of 0.9431 and an F1 score of 0.9389; the Viral Pneumonia category received an F1 score of 0.9504. Table 1 details the test reports of each type of chest radiograph under different models. From the classic deep learning model analysis, for COVID19 data set, the deeper the network layer, the worse the effect of the model. For example, the test results of the Densenet201 model only get good prediction results in a few categories. GoogLeNet obtains the best results in the classic deep learning network, but compared to the MFDNN model, the traditional deep learning model does not achieve the best test results.

**Table 1.** Densenet201, ResNet50, VGG19, GoogLeNet, MFDNN classification technical report.

|  |  | Densenet201 | ResNet50 | VGG19 | GoogLeNet | MFDNN |
| --- | --- | --- | --- | --- | --- | --- |
| COVID | Precision | 0.9272 | 0.9473 | 0.9532 | **0.9572** | 0.9369 |
|  | Recall | 0.8105 | 0.87 | 0.7331 | 0.8976 | **0.9447** |
|  | F1 score | 0.8649 | 0.907 | 0.8288 | 0.9264 | **0.9358** |
| Lung_Opacity | Precision | 0.7198 | 0.801 | 0.7968 | 0.854 | **0.9144** |
|  | Recall | **0.9359** | 0.9143 | 0.8611 | 0.9002 | 0.9068 |
|  | F1 score | 0.8137 | 0.8539 | 0.8277 | 0.8765 | **0.9106** |
| Normal | Precision | **0.941** | 0.9302 | 0.8627 | 0.9266 | 0.9348 |
|  | Recall | 0.8057 | 0.89 | 0.9001 | 0.923 | **0.9431** |
|  | F1 score | 0.8681 | 0.9097 | 0.881 | 0.9248 | **0.9389** |
| Viral Pneumonia | Precision | 0.9007 | **0.9675** | 0.956 | 0.9611 | 0.9257 |
|  | Recall | **0.9777** | 0.8847 | 0.8885 | 0.9182 | 0.9765 |
|  | F1 score | 0.9376 | 0.9242 | 0.921 | 0.9392 | **0.9504** |
| Test Accuracy |  | 85.44% | 89.32% | 85.99% | 91.13% | **93.19%** |

Secondly, Figure 3 describes the confusion matrix of each model prediction test set. This can give us a rough idea of how all images are classified and where most misclassifications occur. It can be seen from the figure that the probability of the prediction error of the Normal class is greater than the probability of the prediction error of the other classes. This shows that the up-sampling method embedded in the algorithm has a positive effect. It makes the model not biased to ignore infected patients during the detection process.

**Figure 3.**
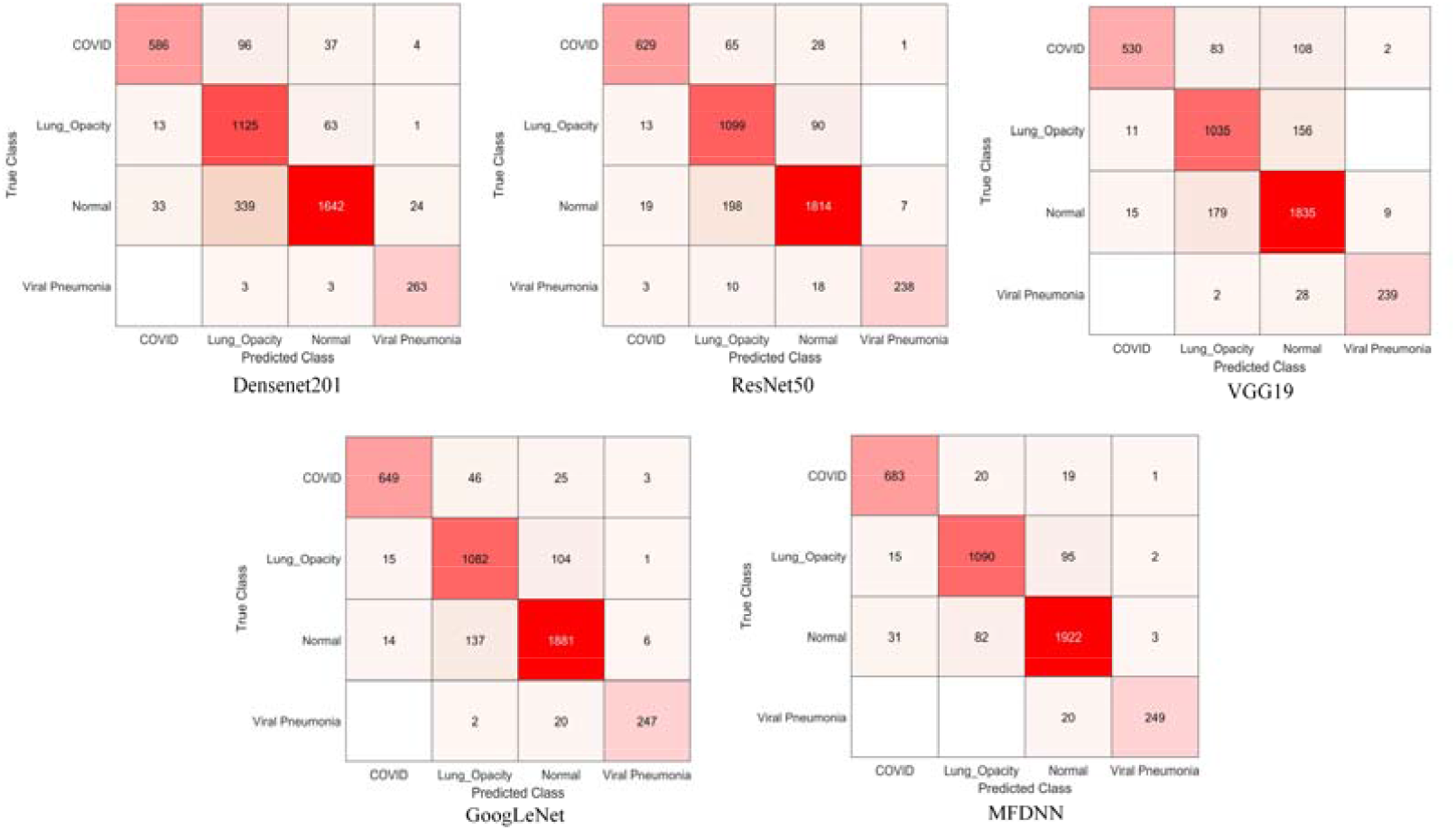
Confusion matrix of Densenet201, ResNet50, VGG19, GoogLeNet, MFDNN model.

### 4.2. Discussion

Most of the common COVID-19 screenings are based on the existing public database. We found that most experiments are based on two classifications (COVID19, Normal) and three classifications (Normal, COVID-19 positive, Viral Pneumonia). The accuracy of the common Deep CNN, Densenet and VGG methods remains below 90% [27] [28] [29]. However, in the actual screening process, the penetration rate of X-rays will image the imaging of chest features. Therefore, adding the Lung Opacity (Non-COVID lung infection) class is beneficial to improve the accuracy of screening for COVID19 infections. Compared with the existing references, the CoroDet model has a classification accuracy of 91.2% on the four-category COVID19 dataset [30]. The accuracy of the MFDNN model proposed in this experiment is 93.19%, which is 1.91% higher than the CoroDet model. Chest X-rays play a key role in screening for COVID19 infections. However, CXI also have some disadvantages. For example, after the rays pass through the chest, the lung characteristics of the infected person are not obvious. Secondly, the radiation of the chest radiograph is relatively large, which can cause certain harm to the human body.

## 5. Conclusion

This paper proposes an MFDNN algorithm to screen people infected with COVID-19. The algorithm integrates data oversampling technology and a multi-channel feature deep neural network model to carry out the training process in an end-to-end manner. In the experiment, we used the publicly available CXI database to train the model. First, by comparing with traditional deep learning models (such as Densenet201, ResNet50, VGG19, GoogLeNet), the MFDNN model obtains an average test accuracy of 93.19% in all data. In each type of detection, most of the experiments are precision, recall, F1 Score is also better than traditional deep learning networks. Secondly, comparing the latest CoroDet model, the MFDNN algorithm is 1.91% higher than the CoroDet model in the four-classification experiment of COVID19 infected persons. However, the limitation of this experiment is mainly in the disadvantages of X-rays. For opaque images of the lungs, RT-PCR is needed to assist in the screening of COVID19 infections.

## Data Availability

please see this URL

https://github.com/panliangrui/covid19

## Acknowledgments

This work was supported by National Key R&D Program of China 2017YFB0202602, 2018YFC0910405, 2017YFC1311003, 2016YFC1302500, 2016YFB0200400, 2017YFB0202104; NSFC Grants U19A2067, 61772543, U1435222, 61625202, 61272056; Science Foundation for Distinguished Young Scholars of Hunan Province (2020JJ2009); Science Foundation of Changsha kq2004010; JZ20195242029, JH20199142034, Z202069420652; The Funds of Peng Cheng Lab, State Key Laboratory of Chemo/Biosensing and Chemometrics; the Fundamental Research Funds for the Central Universities, and Guangdong Provincial Department of Science and Technology under grant No. 2016B090918122.

